# Dysautonomia in hypermobile Ehlers-Danlos syndrome and hypermobility spectrum disorders is associated with exercise intolerance and cardiac atrophy

**DOI:** 10.1101/2021.02.08.21251338

**Authors:** Tania Ruiz Maya, Veronica Fettig, Lakshmi Mehta, Bruce D. Gelb, Amy R. Kontorovich

**Affiliations:** Zena and Michael A. Wiener Cardiovascular Institute, Icahn School of Medicine at Mount Sinai, New York, NY; Department of Genetics and Genomic Sciences, Icahn School of Medicine at Mount Sinai, New York, NY; Department of Pediatrics, Icahn School of Medicine at Mount Sinai, New York, NY; The Mindich Child Health and Development Institute Icahn School of Medicine at Mount Sinai, New York, NY

**Keywords:** hypermobile Ehlers-Danlos syndrome, hypermobility spectrum disorders, dysautonomia, exercise, echocardiography

## Abstract

Dysautonomia is a recognized manifestation in patients with joint hypermobility (JH) disorders. Symptoms can be highly debilitating and commonly include physical deconditioning and poor aerobic fitness. In this study, the prevalence of dysautonomia, range of associated symptoms, patient-reported physical activity levels and echocardiographic features were assessed retrospectively in a cohort of 144 patients (94% female) with hypermobile Ehlers-Danlos syndrome (hEDS) or hypermobility spectrum disorder (HSD). Echocardiographic parameters of LV size and function were compared between patients with and without dysautonomia, as well as to reported values from healthy controls. Dysautonomia was identified in 71% of female and 56% of male subjects and was associated with a high burden of symptomatology, most commonly exercise intolerance (78%). Exercise capacity was limited by dysautonomia, often postural symptoms, in half of all patients. We observed a reduction in physical activity following the onset or significant flare of hEDS/HSD, most strikingly noting the proportion of dysautonomic patients with sedentary lifestyle, which increased from 39% to >80%. JH-related dysautonomia was associated with smaller cardiac chamber sizes, consistent with previous reports in positional orthostatic tachycardia syndrome. Dysautonomia is highly prevalent in patients with hEDS/HSD, exercise intolerance is a key feature and leads to drastic decline in physical activity. Unfavorable cardiac geometry may underlie dysautonomia symptoms and may be due to cardiac atrophy in the setting of aerobic deconditioning.

## Introduction

Symptomatic joint hypermobility (JH) often exists as part of a broader syndromic diagnosis, widely believed to be due to a set of autosomal dominant disorders of connective tissue including the Ehlers-Danlos syndromes (EDS) (Castori et al., 2017). These syndromes, manifesting cardinally as joint instability, chronic joint trauma, chronic musculoskeletal pain and disturbed proprioception, are estimated to afflict 10 million in the United States and 255 million worldwide but are widely under-recognized and underdiagnosed (Kumar & Lenert, 2017; Tinkle et al., 2017).

Systemic symptoms are common in patients with JH-related disorders and include palpitations, atypical chest discomfort, fatigue, dizziness/lightheadedness, syncope, exercise intolerance, dependent acrocyanosis, peripheral vasoconstriction, and impaired thermoregulation (A. Hakim et al., 2017; A. J. Hakim, Cherkas, Grahame, Spector, & MacGregor, 2004). Tachycardia and hypotension are long recognized complications, typically presenting as orthostatic intolerance (OI), orthostatic hypotension (OH), positional orthostatic tachycardia syndrome (POTS) and neurally-mediated hypotension (NMH)(A. Hakim et al., 2017), a collection of diagnoses falling broadly under the umbrella of dysautonomia (Robertson, 1999). Systemic dysautonomia has been tightly linked to EDS since 1999 when Rowe *et al*. showed that all of twelve pediatric EDS patients had either POTS or NMH (Rowe et al., 1999). Dysautonomia symptoms have been reported at high rates (up to 78%) in other adult JH and EDS cohorts (Gazit, Nahir, Grahame, & Jacob, 2003; A. Hakim et al., 2017), with POTS as the most prevalent autonomic profile (De Wandele et al., 2014). Proposed mechanisms include sympathetic neurogenic dysfunction, vascular connective tissue laxity, sudomotor impairment and use of vasoactive medications (De Wandele et al., 2014; Wallman, Weinberg, & Hohler, 2014).

Physical deconditioning and poor aerobic fitness are commonly cited in patients with JH-related disorders, often with dysautonomia symptom onset following a prolonged period of reduced physical activity (*e*.*g*., following joint injury)(A. Hakim et al., 2017). A relationship between deconditioning and dysautonomia has previously been established. Fu *et al*. found that the marked tachycardia during orthostasis in patients with POTS was attributable to small cardiac size coupled with reduced blood volume, engendering the moniker “Grinch Syndrome” after the infamous Dr. Seuss character whose heart was “two sizes too small”(Fu et al., 2010). This finding, in contrast to the observed left ventricular (LV) hypertrophy and increased cardiac volumes seen in well-conditioned athletes (Fagard, 2003; Pelliccia, Maron, Culasso, Spataro, & Caselli, 1996), led to a proposed association between POTS and aerobic deconditioning. In further support, aerobic reconditioning therapy through exercise improves or resolves POTS in most patients (George et al., 2016; Winker et al., 2005) while also significantly increasing LV mass and blood volumes (Fu et al., 2010; Fu et al., 2011). It is unknown whether small heart size explains the broad dysautonomia accompanying JH-related disorders, or whether aerobic training can offer a similar benefit in this population.

In 2017, nosology for JH disorders were updated to delineate between two forms: 1) hypermobile EDS (hEDS, previously known as EDS type III and EDS hypermobility type) and the hypermobility spectrum disorders (HSD). The HSDs, further sub-grouped as “generalized” (G-HSD), “peripheral” (P-HSD), “localized” (L-HSD) or “historical” (H-HSD), are thusly defined as distinguishable from hEDS by virtue of a phenotype restricted to the musculoskeletal system and not involving other systemic manifestations (Castori et al., 2017), although clinical severity can be similar (Copetti et al., 2019). In the era of this contemporary hEDS/HSD nosology, there is an incomplete descriptive inventory of the prevalence and spectrum of associated dysautonomia. In this single-center retrospective cohort study of 144 patients with JH-related disorders, we report the prevalence of dysautonomia in hEDS and HSDs, the comprehensive range of associated symptomatology, physical activity levels before and after the onset of hEDS/HSD and cardiac morphologic profiles that may contribute to dysautonomia in these groups.

## Methods

Included subjects were those seen in a multidisciplinary Cardiovascular Genetics Program for evaluation of a JH-related disorder between January 2017 and January 2020, referred by cardiologists, primary care providers, rheumatologists, orthopedists, pain management specialists, neurologists or self-referred and evaluated by a study investigator (A.R.K, B.D.G. or L.M.). The study was conducted under the approval of Mount Sinai’s Institutional Review Board (GCO # 19-0883 ISMMS). Electronic health records (EHR) were retrospectively reviewed for demographic data and diagnoses of hEDS or HSD documented by a study investigator within the progress note linked to the in-person clinical evaluation. The diagnosis of hEDS or HSD was determined according to the 2017 consensus guidelines including degree of hypermobility based on the Beighton score, presence of systemic and echocardiographic features, family history and exclusion of alternative genetic or rheumatologic diagnoses (Castori et al., 2017). Ascertainment of dysautonomia diagnosis was made through review of progress note documentation. Patient-reported symptoms previously described in association with dysautonomia were systematically collected, including: (a) resting sinus tachycardia or palpitations reported in a single category representing a rapid heart rate, (b) chronic fatigue, (c) chest pain or chest discomfort either at rest and during exertion and of any quality, (d) hypotension from historical data provided by patient or objective data within the EHR, (e) dyspnea, (f) near syncope, or true syncope, (g) dizziness related to postural changes reported with onset by changing from a supine to upright position or with maintaining upright posture, (h) cognitive complaints including concentration impairment (most frequent), “brain fog,” word finding difficulties and poor memory, (i) lower extremity edema and/or discoloration often described as “blood pooling” after standing for relatively short periods of time, (j) temperature intolerance or dysregulation either explicitly reported by patients or as complaints of cold, dusky extremities and/or heat as a trigger for other dysautonomia-related symptoms such as near-syncope, (k) gastrointestinal symptoms including chronic or episodic constipation, nausea and/or diarrhea, (l) complaints of underactive and/or overactive bladder symptoms. Next, we systematically extracted physical activities reported before the onset or flare of hEDS/HSD symptoms (“baseline”) and at time of the initial evaluation (“current”). Patient exercise levels were categorized as either: 1) none, 2) light, 3) moderate or 4) vigorous based on highest intensity activity reported and as per established guidelines and recommendations (Levine et al., 2015; Maron et al., 2004; Metkus, Baughman, & Thompson, 2010) (Supplemental Table 1).

**Table 1.** **Prevalence of Dysautonomia in hypermobile Ehlers-Danlos syndrome and Hypermobility Spectrum Disorders.**

| **Diagnosis** | **Total** | **Total** | **Dysautonomia** | **Dysautonomia** |
| --- | --- | --- | --- | --- |
|  | **N (%)** | **M/F** | **N (%)** | **M/F** |
| **hEDS†** | 60 (41.7) | 3/57 | 48 (47.5) | 2/46 |
| **G-HSD** | 51 (35.4) | 4/47 | 33 (32.7) | 2/31 |
| **L-HSD** | 17 (11.8) | 1/16 | 9 (8.9) | 1/8 |
| **H-HSD** | 12 (8.3) | 1/11 | 8 (7.9) | 0/8 |
| **A-GJH** | 4 (2.8) | 0/4 | 3 (3.0) | 0/3 |
| **Total** | **144** | **9/135** | **101 (70.13)** | **5/96** |

†One Female to Male transgender patient.
A- GJH- Asymptomatic Generalized Joint Hypermobility
G- HSD- Generalized-Hypermobility Spectrum Disorder
hEDS-Hypermobile Ehlers-Danlos syndrome
H- HSD- Historical-Hypermobility Spectrum Disorder
L- HSD- Localized-Hypermobility Spectrum Disorder

2-D echocardiographic data was collected from clinical reports where available, to record the following: LV ejection fraction (EF), LV internal diastolic diameter (LVIDd) and LV end-diastolic volume (LVEDV) indexed to body surface area (LVEDV/BSA). LV mass was calculated by linear method as recommended by the American Society of Echocardiography (ASE) with the Devereux formula: LV mass = 0.8 × {1.04 [(LVID + PWT + IVSWT)^3^ −(LVID)^3^]} + 0.6, with all measurements obtained in diastole (Lang et al., 2015) and indexed to BSA (LV mass/BSA). Means of each parameter were compared for hEDS/HSD patients with vs. without dysautonomia. Mean LVIDd and LV mass/BSA were compared with previously published gender-specific mean reference normal values derived from non-hypertensive heart-healthy controls (Cuspidi et al., 2012).

Descriptive statistics for categorical variables were reported as frequency and percentages, continuous variables were reported as median with first and third quartiles or mean and standard deviation. Statistical analysis was performed using SPSS version 25 (IBM Corp., Armonk, NY) and Microsoft Excel. Fisher’s exact test (two-tailed) was used to compare rates of dysautonomia between hEDS and HSD cohorts as well as to compare proportion of sub-cohorts having echocardiographic measurements below a reference normal. The Student’s t-test (2-tailed, heteroscedastic or homoscedastic after calculation of variance using the F-test) was used to compare the distribution of echocardiographic values in hEDS/HSD subjects with vs. without dysautonomia. The one sample t-test was used to compare calculated mean echocardiographic values for the cohorts against published reference normal mean values. P<0.05 was considered as statistically significant for the Fisher’s exact test, Student’s t-tests and one sample t-test.

## Results

During the two-year study period, 144 patients were evaluated for and diagnosed with a JH-related disorder, 60 (42%) with hEDS and 84 (58%) with a HSD (Table 1). The median age was 31 years (interquartile range 25-42) and 94% were female (Table 2). In total, 101 (96 female, 5 male) patients (70%) were diagnosed with dysautonomia (Table 1), more commonly in hEDS vs. HSD (48, 80% vs. 53, 63% of the respective primary diagnoses, p=0.04). The most frequently reported dysautonomia-associated symptom was exercise intolerance (78%), followed by fatigue, dizziness, gastrointestinal (GI) symptoms and palpitations (Figure 1a). Most (93%) patients reported more than one dysautonomia-related symptom with over half reporting more than five (Figure 1b). Because exercise intolerance was so common in this cohort and hEDS/HSD patients often suffer with chronic musculoskeletal pain, we evaluated the self-reported explanation for limitation to exercise. Importantly, dysautonomia-related symptoms (50%) were implicated as frequently as pain (50%) in explaining patients’ inability to exercise at their baseline (Figure 2a). Of the dysautonomia symptoms restricting exercise, postural symptoms were the most common complaint (62%, Figure 2b).

**Table 2.** **Patient Characteristics**

| **Variables** | **Total**<br>**(N=144)** | **Dysautonomia**<br>**(N=101)** | **No Dysautonomia**<br>**(N=43)** | **p-**<br>**value** |
| --- | --- | --- | --- | --- |
| **Age (y)** | 31 [25-42] | 33 [24-42.5] | 30 [26-42] | 0.7 |
| **Sex (female)** | 135 (93.8%) | 96 (95.0%) | 39 (90.7%) | 0.5 |
| **Height (cm)** | 165.1 [160.3-172.7] | 165.1 [161.0-172.6] | 165.1 [160.0-173] | 1 |
| **Weight (kg)** | 64.1 [56.8-73.7] | 65.0 [56.5-74.0] | 63.5 [57.2-72.0] | 1 |
| **Body mass index (kg/m<sup>2</sup>)** | 22.9 [20.4-26.4] | 22.9 [20.3-27.0] | 22.9 [21.4-25.8] | 0.8 |
| **Body surface area (m<sup>2</sup>)** | 1.72 [1.60-1.83] | 1.72 [1.61-1.83] | 1.69 [1.56-1.85] | 0.8 |
| **Left ventricular ejection**<br>**fraction†** | 62.5 [60-65] | 62.5 [60-66] | 60.5 [60-65] | 0.6 |

Continuous variables expressed as a median [25th, 75th percentiles].
†Echocardiographic data available for 108 (75%) of patients, 74/101 with dysautonomia and 34/43 without dysautonomia.

**Figure 1.**
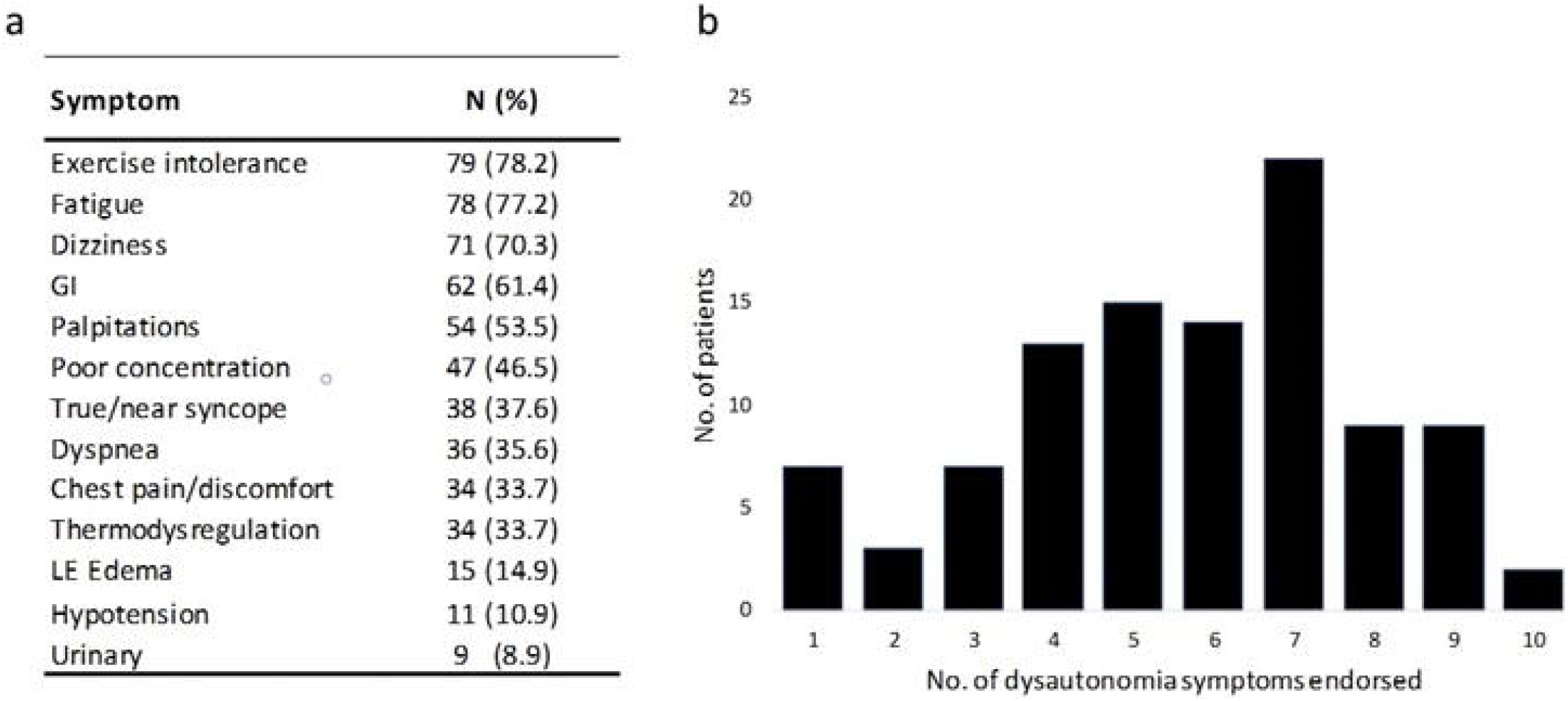
Prevalence and extent of symptoms in hypermobile Ehlers-Danlos syndrome and hypermobility spectrum disorder patients with dysautonomia. a) More than half of hEDS/HSD patients with dysautonomia endorsed palpitations and >75% endorsed exercise intolerance. Data expressed as total number and percentage of dysautonomia patients; GI: gastrointestinal symptoms including nausea, vomiting/regurgitation, constipation, diarrhea, and/or abdominal pain; LE: lower extremity. b) Multiple symptomatology was the rule, with >50% of patients having more than five dysautonomia symptoms.

**Figure 2.**
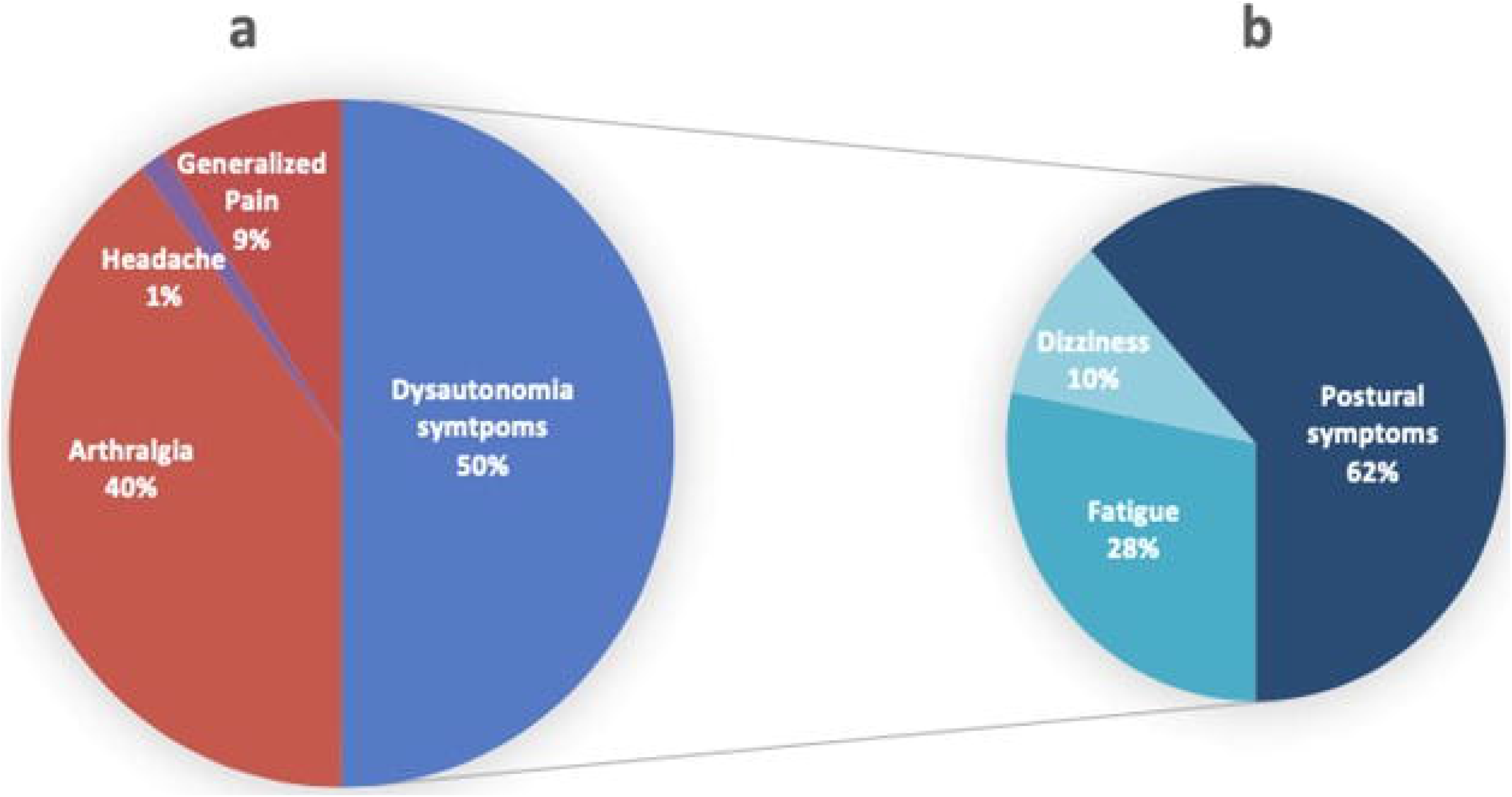
Self-reported limitation to achieving pre-hypermobility Ehlers-Danlos syndrome or pre-hypermobility spectrum disorder onset/flare exercise level. a) Barriers to exercise were either physical pain related to the underlying joint hypermobility-related disorder or dysautonomia symptoms. b) Of those reporting dysautonomia symptoms as their major limitation to achieving pre-joint hypermobility-related disorder onset levels of exercise, most endorsed postural symptoms.

At the time of hEDS/HSD initial evaluation, >75% of subjects reported a sedentary lifestyle, meeting the self-reported physical activity level designation of “none” (N=49, 34%) or “light” (N=60, 42%); only 35 (24%) reported exercising at a “moderate” or “vigorous” level. In contrast, prior to the onset or significant flare of hEDS/HSD, 60% of subjects exercised at a “moderate” (N=61, 42%) or “vigorous” (N=26, 18%) level; only 19 (13%) of patients performed no formal exercise prior to hEDS/HSD onset/flare (Supplemental Figure 1). While 62 (61%) of dysautonomia patients were moderate/vigorous exercisers at baseline, this number dropped to 20 (20%) following hEDS/HSD onset/flare (Supplemental Figure 1a). The difference was less dramatic among non-dysautonomia patients (Supplemental Figure 1b), where 25 (58%) vs. 15 (35%) were moderate/vigorous exercisers at baseline compared to after hEDS/HSD onset/flare. Notably, the number of patients reporting no regular physical activity (“none”) at baseline was 16 (16%) among dysautonomia and 3 (7%) among non-dysautonomia patients. At the time of hEDS/HSD evaluation, 21% of patients reported engaging in a formal physical therapy (PT) program (focused on musculoskeletal symptoms); 3% of these reported significant improvement of arthralgia and improved exercise tolerance with PT, but none could perform at their baseline exercise level due to persistent dysautonomia-related symptoms.

LV chamber sizes were smaller overall for hEDS/HSD patients with dysautonomia compared to those without (Table 3). For both males and females, mean LVIDd were lower in the dysautonomia vs. non-dysautonomia groups, although this only reached statistical significance for females. For males, LVEDV/BSA was significantly lower, while for females there was a non-significant lower trend. Indexed LV mass trended lower in dysautonomia groups but differences were not statistically significant.

**Table 3.**
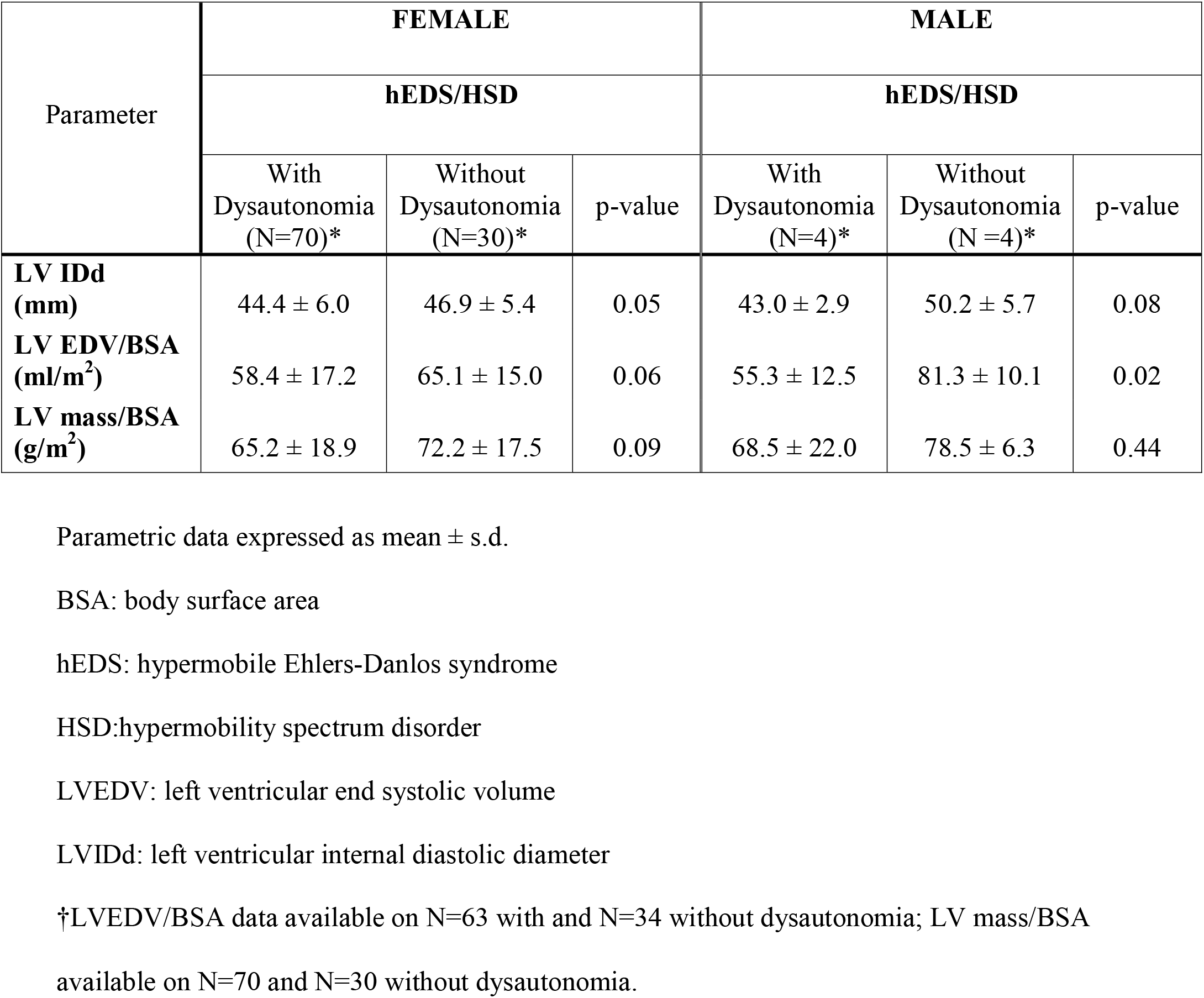
**Comparison of 2D echocardiographic dimensions for parameters of left ventricular chamber size by gender and dysautonomia status.**

| Parameter | FEMALE |  |  | MALE |  |  |
| --- | --- | --- | --- | --- | --- | --- |
|  | hEDS/HSD |  |  | hEDS/HSD |  |  |
|  | With Dysautonomia (N=70)* | Without Dysautonomia (N=30)* | p-value | With Dysautonomia (N=4)* | Without Dysautonomia (N =4)* | p-value |
| **LV IDd (mm)** | 44.4 ± 6.0 | 46.9 ± 5.4 | 0.05 | 43.0 ± 2.9 | 50.2 ± 5.7 | 0.08 |
| **LV EDV/BSA (ml/m<sup>2</sup>)** | 58.4 ± 17.2 | 65.1 ± 15.0 | 0.06 | 55.3 ± 12.5 | 81.3 ± 10.1 | 0.02 |
| **LV mass/BSA (g/m<sup>2</sup>)** | 65.2 ± 18.9 | 72.2 ± 17.5 | 0.09 | 68.5 ± 22.0 | 78.5 ± 6.3 | 0.44 |

Parametric data expressed as mean ± s.d.
BSA: body surface area
hEDS: hypermobile Ehlers-Danlos syndrome
HSD:hypermobility spectrum disorder
LVEDV: left ventricular end systolic volume
LVIDd: left ventricular internal diastolic diameter
†LVEDV/BSA data available on N=63 with and N=34 without dysautonomia; LV mass/BSA available on N=70 and N=30 without dysautonomia.

When compared to a heart-healthy reference non-hypertensive control population (Cuspidi et al., 2012), LVIDd were smaller for dysautonomic males (43 mm vs. 50 mm, p=0.02) and females (44 mm vs. 46 mm, p=0.03) but not those without dysautonomia (50 mm vs. 50 mm for males, p=0.95; 47 mm vs. 46 mm for females, p=0.36) (Supplemental Table 2). Mean indexed LV mass was significantly lower for dysautonomic females (65 g/m^2^ vs. 72 g/m^2^, p<0.01) but not non-dysautonomic females (72 mL vs. 72 mL, p=0.98) or any males compared to the respective reference means.

## Discussion

Hypermobility disorders are strikingly common but rarely diagnosed (Kumar & Lenert, 2017). A conservative estimate is that hEDS/HSD affects 3% of the population (Mulvey et al., 2013), making these disorders >100 times more prevalent than Marfan syndrome. Dysautonomia is considered a common comorbidity, but the prevalence in hEDS and HSDs has not been defined separately in the wake of contemporary nosology. Of the 144 patients in this study with one of these disorders, 70% had concomitant dysautonomia. Notably, the rate of dysautonomia was higher in females (71%) than males (56%), although the small number of males in this study precludes meaningful statistical comparisons. As with JH disorders, symptom burden is higher in dysautonomic women, often those of child-bearing age (Garland, Raj, Black, Harris, & Robertson, 2007). Patients with JH-related disorders (Scheper et al., 2016) and those with POTS (Raj, 2013) report low health-related quality of life and are often unable to go to school, work or participate in recreational activities. As we found in this study and has been shown previously (Wallman et al., 2014), these disorders are often found in tandem, putting this population of largely young women at risk for significant morbidity and decreased quality of life over many years.

In the present study, dysautonomia was common among both hEDS and HSD cohorts, indicating a likely shared pathophysiologic mechanism, however, the prevalence was significantly higher in hEDS patients. Still, our results show that dysautonomia cannot be used as a discriminator between these diagnoses. The dysautonomic symptomatology observed in our hEDS/HSD cohort was wide and can have a considerable impact on quality of life. Indeed, in our cohort, >90% of patients with dysautonomia had three or more related symptoms. Exercise intolerance was notably common among JH-related dysautonomia patients, occurring in >75%. While it is possible that exercise intolerance for some was due to physical limitations and pain from underlying hEDS/HSD, most individuals reported this as part of a collection of other classic dysautonomia symptoms, and only a handful cited exercise intolerance as their only symptom, fewer in dysautonomia vs. non-dysautonomia patients (2, 2% vs. 3, 7%). This particular scenario inevitably invokes a “chicken or egg” causality quagmire: does exercise intolerance beget dysautonomia or vice versa? In a recent review of POTS where EDS and JH were recognized as frequently associated conditions, the most common demographic noted was young, previously active women with an identifiable event preceding symptom onset and resulting in bed rest or withdrawal of physical activity (Bryarly, Phillips, Fu, Vernino, & Levine, 2019). Determining the natural history of exercise intolerance in JH-related dysautonomia was outside the scope of the present study but warrants further examination since physical therapy exercises are the cornerstone of treatment for hEDS/HSD.

Dysautonomic mechanisms in POTS patients are postulated to relate to aerobic deconditioning as evidenced by the observation of lower physical performance due to lower stroke volumes during exercise (Fu et al., 2011; Shibata et al., 2012) and exercise training in POTS expands plasma volume resulting in increases in cardiac size and mass as well as improved orthostatic tolerance (George et al., 2016). These mechanisms have not been explored more broadly in dysautonomia patients, especially those with hEDS/HSD. However, our hEDS/HSD dysautonomia cohort displays many similarities to POTS patients, including overlap in symptoms, especially exercise intolerance and deconditioning (Fu et al., 2010). Furthermore, a significant proportion of patients in our cohort reporting high levels of physical activity and exercise at baseline endorsed a decline to none/light activity following a period of inactivity due to acute illness (*e*.*g*., motor vehicle accident, tendon tear, flare of gastrointestinal symptoms). In the POTS literature, a history of cardiovascular dysfunction following a prolonged period of reduced activity is common (A. Hakim et al., 2017). This lends further credence to the hypothesis that aerobic deconditioning may underlie dysautonomic symptoms in hEDS/HSD. Considering that patients with hEDS/HSD suffer flares of musculoskeletal pain and frequent joint and soft-tissue injuries (Castori, 2016), such individuals would be at particularly high risk. Additionally, we found that patients without dysautonomia had overall higher exercise levels before and after the onset or flare of their hEDS/HSD compared to those with dysautonomia, with 93% vs. 84% reporting engaging in some formal physical activity and only 23% vs. 39% dropping down to no exercise thereafter. This raises the possibility that continued basal exercise throughout hEDS/HSD flares may be protective against the development of dysautonomic symptomatology. Further prospective case-control studies are needed to investigate this prospect.

The mechanisms underlying autonomic dysfunction in hEDS/HSD remain unclear and this study was not designed to define them, but are suggested to include low blood pressure, increased peripheral venous dilation and blood pooling, low circulating blood volume, elevated circulating catecholamines and excess systemic levels of histamine (A. Hakim et al., 2017).

There is evidence supporting some of these mechanisms in hEDS such as increased aortic wall compliance (Handler, Child, Light, & Dorrance, 1985), abnormal connective tissue in dependent blood vessels causing excessive distention in response to ordinary hydrostatic pressures (Rowe et al., 1999), neuropathy, connective tissue laxity, alpha and beta adrenergic hyper-responsiveness (De Wandele et al., 2014; Mathias et al., 2011), as well as potentially pathogenic adrenergic, muscarinic and other neural autoantibodies (A. Hakim et al., 2017). Proposed mechanisms for autonomic dysfunction in patients with POTS specifically include a small cardiac chamber size and low plasma volumes (Fu et al., 2010), and considering the symptom overlap, it is reasonable to consider that these features may be part of the hEDS/HSD dysautonomia phenotype as well. Our findings of smaller LV dimensions in hEDS/HSD patients with vs. without dysautonomia supports this theory, although the differences were only statistically significant in females when looking at LVIDd and in males when looking at indexed LVEDV. Still, for a retrospective study with non-standardization of echocardiograms (see Limitations below), this is an intriguing result that warrants further study using a standardized prospective design.

## Conclusion

In summary, patients with either hEDS/HSD or dysautonomia are at heightened risk for substantial morbidity and impaired quality of life, but carrying both diagnoses imposes an even greater symptomatic burden and disproportionately affects young women. hEDS and HSD lead to exercise intolerance and avoidance, likely further potentiating symptoms and sparking an unfortunate cycle with sedentary lifestyle as a cause and consequence. Cardiac atrophy may be a feature of this phenomenon and aerobic reconditioning to more favorably remodel the cardiac geometry is a potentially promising intervention that deserves further investigation.

## Data Availability

The data that support the findings of this study are available from the corresponding author upon reasonable request.

## Authorship confirmation statement

All persons who meet authorship criteria are listed as authors, and all authors certify that they have participated sufficiently in the work to take public responsibility for the content, including participation in the concept, design, analysis, writing, or revision of the manuscript.

