## Supplemental Tables for "Dysautonomia in hypermobile Ehlers-Danlos syndrome and hypermobility spectrum disorders is associated with exercise intolerance and cardiac atrophy"

**Supplemental Table 1. Exercise level categories based on highest tolerated patient-reported physical activity***

| None | Light | Moderate | Vigorous |
| --- | --- | --- | --- |
| No formal exercise beyond activities of daily living and walking to and from work at a regular pace. | Formal walking program  Yoga  Pilates  Gardening  Yard work  Physical therapy  Golf  Badminton  Croquet  Shuffleboard  Bowling  Ping-pong | Brisk walking  Recreational sports  Stair climbing  Elliptical machine  Dancing  Light swimming  Cycling on flat ground  Hiking  Doubles tennis | Jogging  Running  Swimming  Singles tennis  Full court basketball  Resistance training  Shoveling snow  Cross country skiing  Mountain biking  Calisthenics  Rowing |

*Categorizations adapted from Maron et al,^19^ Levine et al,^20^ and Metkus et al.^21^

**Supplemental Table 2. Left ventricular sizes in hypermobile Ehlers-Danlos syndrome and hypermobility spectrum disorder patients with and without dysautonomia compared to healthy reference controls.**

| Parameter | **FEMALE** | | | | |  | **MALE** | | | |
| --- | --- | --- | --- | --- | --- | --- | --- | --- | --- | --- |
|  |  | **hEDS/HSD** | | | |  | **hEDS/HSD** | | | |
|  | Reference normal | with dysautonomia |  | without  dysautonomia | p | Reference normal | with dysautonomia |  | without  dysautonomia | p |
|  |  |  | p |  |  |  |  | p |  |  |
|  |  | N=70^*^ |  | N=30^*^ |  |  | N=4* |  | N=4* |  |
| **LVIDd**  **(mm)** | 46.0^†^ | 44.4 ± 6.0 | 0.03 | 46.9 ± 5.4 | 0.36 | 50.0^†^ | 43.0 ± 2.9 | 0.02 | 50.2 ± 5.7 | 0.95 |
| **LV mass/BSA**  **(g/m2)** | 72.3^†^ | 65.2 ± 18.9 | <0.01 | 72.2 ± 17.5 | 0.98 | 84.5^†^ | 68.5 ± 22.0 | 0.24 | 78.5 ± 6.3 | 0.15 |

Parametric data expressed as mean ± s.d.

BSA: body surface area

LVIDd: left ventricular internal diastolic diameter

LV mass/BSA data available on N=70 with and N=30 without dysautonomia

^†^Reference normal from Cesare et al (2012). Journal of Hypertension 30(5):997-1003
